# Effectiveness of convalescent plasma therapy in COVID-19 patients with haematological malignancies

**DOI:** 10.1101/2022.04.06.22273542

**Authors:** Sapha Shibeeb, Ilham Ajaj, Hadeel Al-Jighefee, Atiyeh Abdallah

## Abstract

**Background:** Immunocompromised patients, including those with haematological malignancies, are among the high-risk group to develop severe coronavirus disease 2019 (COVID-19) complications. The effectiveness of passive immunotherapy with convalescent plasma (CP) on such patients diagnosed with COVID-19 has not been reviewed. Therefore, the aim of this review was to systematically appraise the current evidence for the efficacy of this therapy in haematological malignancies patients with COVID-19 infection.

**Methods:** A comprehensive search was conducted up-to October 2021, using four databases: PubMed, Web of Science, Science Direct, and Scopus. Two reviewers independently assessed the quality of the included studies. Data collection analysis were performed using Microsoft Excel 365 and GraphPad Prism software.

**Results:** 17 studies met the inclusion criteria; these records included 258 COVID-19 patients with haematological malignancies and treated with CP therapy (CPT). The main findings from the reviewed data suggests CPT may be associated with improved clinical outcomes including (a) higher survival rate, (b) improved SARS-CoV-2 clearance and presence of detectable anti-SARS-CoV-2 antibodies post CP transfusion, (c) improved hospital discharge time, and recovery after 1 month of CP therapy. Furthermore, treatment with convalescent plasma was not associated with development of adverse events.

**Conclusion:** Owing to its safety and beneficial effects in improving clinical outcomes, CPT appears to be an effective supportive therapeutic option for haematological malignancy patients infected with COVID-19.

## Introduction

Since the first case report in December 2019, the severe acute respiratory syndrome coronavirus 2 (SARS-CoV-2), the causative agent of the coronavirus disease 2019 (COVID-19), has posed a significant challenge worldwide (1). The clinical manifestation of COVID-19 ranges from having no signs or symptoms (asymptomatic) to severe complications that include thrombosis, septic shock, acute respiratory distress syndrome (ARDS), and cardiac failure (2). Among those who are at high-risk to severe and prolonged disease course, immunocompromised patients and cancer patients (3, 4). Patients with haematological malignancies represent a distinctive subset of those vulnerable to COVID-19 and were shown to be frequently associated with high mortality and COVID-19 complications (5). Due to the underlying disease and cancer treatment, the immune system in these patients becomes impaired making them immunodeficient and prone to infection and severe disease. (2).

CPT is a form of passive immunity where plasma enriched with specific and non-specific humoral innate immunity factors is collected from recovered patients, processed, and transfused into other patients (6). During viral infection, the antibodies are key for virus opsonisation and neutralisation, in addition to the activation of complement and mediation of antibody dependent cellular cytotoxicity. This type of treatment has been previously used to treat other infectious diseases such as Ebola, SARS, Middle East respiratory syncytial virus (MERS), and influenza (2). Currently, COVID-19 has very limited treatment options, where isolation and supportive care are the major ones (7). In August 2020, the United States Food and Drug Administration (US FDA) issued an emergency use authorization (EUA) for COVID-19 convalescent plasma for the treatment of hospitalized patients with COVID-19 (8). As a result, numerous trials have been conducted to assess the effectiveness of CPT in different COVID-19 patient cohorts including those with different disease severity and co-morbidities. The effectiveness of COVID-19 CPT in immunocompromised cancer patients, particularly, those with haematological malignancies has not been systematically reviewed yet. Therefore, we conducted a systematic review on the effectiveness of CPT to treat COVID-19 patients with haematological malignancies.

## Methods

The study’s focus was on haematological malignancies patients with confirmed PCR COVID-19 infection. The intervention was CPT from previously infected COVID-19 patients. The primary outcomes were (i) clinical improvement and (ii) viral clearance. The secondary outcome was adverse events after CPT. The search was conducted up-to October 2021, using four major databases; PubMed, Web of Science, ScienceDirect, and Scopus. The following terms were used; ‘COVID-19’ or ‘SARS-CoV-2’ or Coronavirus’ AND ‘convalescent plasma’ or ‘convalescent plasma therapy’. All articles were extracted and assessed by two independent authors to ensure the quality of the research. All articles were exported to Endnote X9 and all duplicates were removed.

### Inclusion and exclusion criteria

Articles were screened by title and abstracts, the following inclusion criteria were used for eligibility: 1) reported in English 2) clinical trials including, randomized and controlled clinical trials 3) prospective and retrospective comparative cohort studies, case-control studies; cross-sectional studies, case series, and case reports. Studies were excluded if they did not meet the inclusion criteria. In addition, all letters, editorials, systematic reviews, narrative reviews, abstracts, and not full-text articles were excluded. Articles published using same patient cohort were grouped and studies with largest number of patients were used.

### Quality assessment and Risk of Bias Assessment

The risk of bias for all eligible observational studies was assessed according to the Strengthening the Reporting of Observational Studies in Epidemiology (STROBE) reporting guideline (9). The risk was evaluated using a previously published question tool (10), which asked questions regarding (1) Selection criteria of patients: Do all the patients meet inclusion criteria (2) Adequate ascertainment of exposure and the outcome (3) Causality: was follow-up long enough for outcomes to occur? (4) Reporting: were the case(s) described with sufficient detail to allow other investigators to replicate the research or to allow practitioners to make inferences? All studies were scored for each question as: yes (2 stars), partial (1 star), and no (0 star). An overall risk of bias was independently assigned to each eligible study by two authors. No studies rated below 5 were included in the systematic review (Table 1).

**Table 1.**
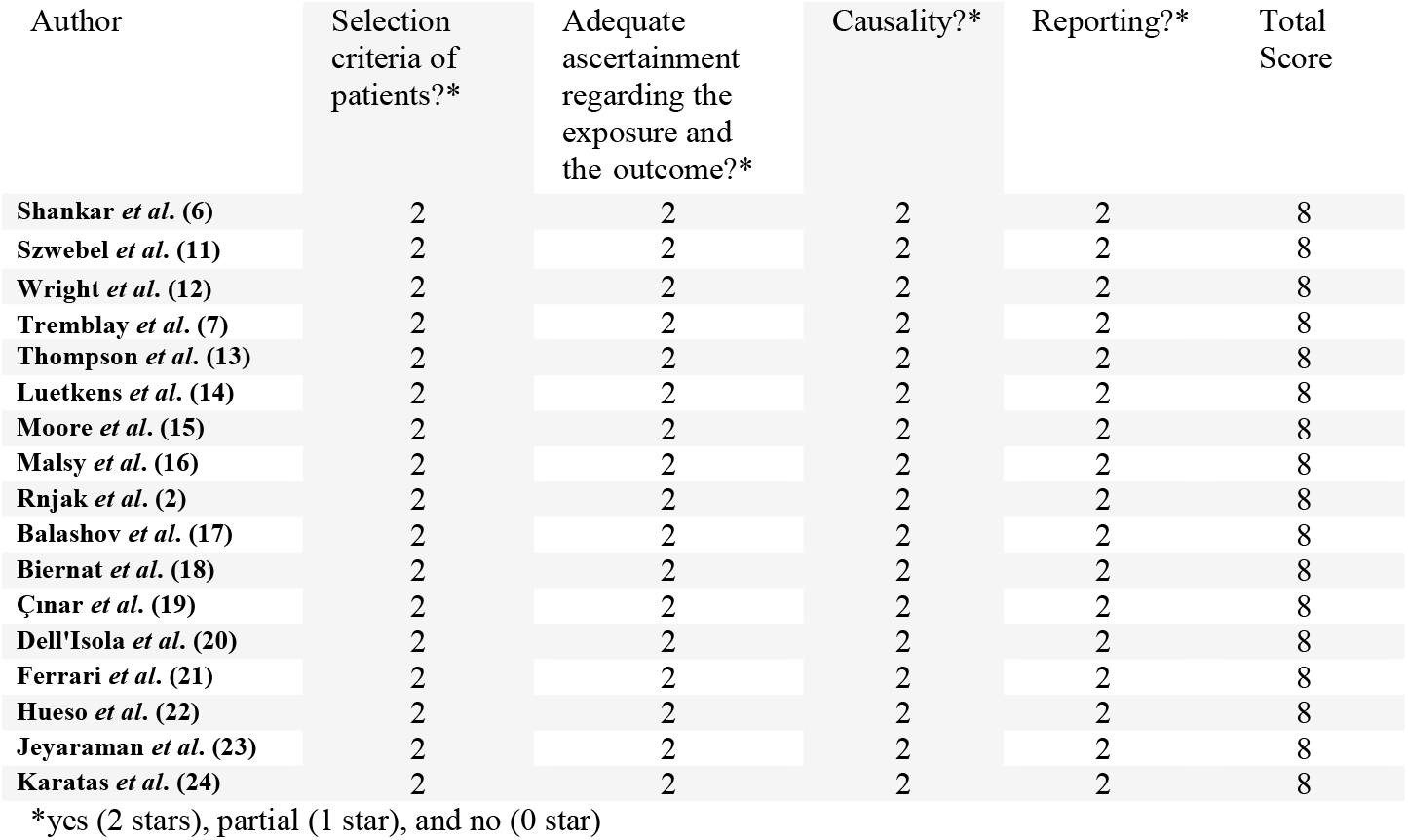
Risk of Bias Assessment

| Author | Selection<br>criteria of<br>patients?* | Adequate<br>ascertainment<br>regarding the<br>exposure and<br>the outcome?* | Causality?* | Reporting?* | Total<br>Score |
| --- | --- | --- | --- | --- | --- |
| **Shankar *et al.* (6)** | 2 | 2 | 2 | 2 | 8 |
| **Szwebel *et al.* (11)** | 2 | 2 | 2 | 2 | 8 |
| **Wright *et al.* (12)** | 2 | 2 | 2 | 2 | 8 |
| **Tremblay *et al.* (7)** | 2 | 2 | 2 | 2 | 8 |
| **Thompson *et al.* (13)** | 2 | 2 | 2 | 2 | 8 |
| **Luetkens *et al.* (14)** | 2 | 2 | 2 | 2 | 8 |
| **Moore *et al.* (15)** | 2 | 2 | 2 | 2 | 8 |
| **Malsy *et al.* (16)** | 2 | 2 | 2 | 2 | 8 |
| **Rnjak *et al.* (2)** | 2 | 2 | 2 | 2 | 8 |
| **Balashov *et al.* (17)** | 2 | 2 | 2 | 2 | 8 |
| **Biernat *et al.* (18)** | 2 | 2 | 2 | 2 | 8 |
| **Çınar *et al.* (19)** | 2 | 2 | 2 | 2 | 8 |
| **Dell'Isola *et al.* (20)** | 2 | 2 | 2 | 2 | 8 |
| **Ferrari *et al.* (21)** | 2 | 2 | 2 | 2 | 8 |
| **Hueso *et al.* (22)** | 2 | 2 | 2 | 2 | 8 |
| **Jeyaraman *et al.* (23)** | 2 | 2 | 2 | 2 | 8 |
| **Karatas *et al.* (24)** | 2 | 2 | 2 | 2 | 8 |

\*yes (2 stars), partial (1 star), and no (0 star)

**Table 2.** Summary of included studies on CPT for COVID-19 patients with haematological malignancies

| Author | Year | Country | Malignancy | Study design | Sample sizes<br>CPT/Non-CPT | Age<br>median<br>(range) | Gender | Transplant |
| --- | --- | --- | --- | --- | --- | --- | --- | --- |
| **Shankar *et al.* (6)** | 2021 | United States | ALL | Case report | 1/0 | 4 years | F |  |
| **Szwebel *et al.* (11)** | 2021 | France | HIV and cancer B-cell lymphoma | Case report | 1/0 | NA | M | ASCT |
| **Wright *et al.* (12)** | 2021 | United States | FL | Case report | 1/0 | 54 years | M |  |
| **Tremblay *et al.* (7)** | 2020 | United States | Hematologic and solid cancer | Case series | 24/0 | 69 years (31-88) | F 41.7%/<br>M 58.3% |  |
| **Thompson *et al.* (13)** | 2021 | United States | Hematologic cancer | Retrospective cohort study | 143/823 | 65 years | NA |  |
| **Luetkens *et al.* (14)** | 2020 | United States | MM | Case report | 1/0 | 72 years | F | 3 ASCT |
| **Moore *et al.***<br>**(15)** | 2020 | United States | NHL | Case report | 1/0 | 63 years | F |  |
| **Malsy *et al.***<br>**(16)** | 2020 | Germany<br>Croatia | FL | Case report | 1/0 | 53 years | F |  |
| **Rnjak *et al.***<br>**(2)** | 2021 | Russia | DLBCL | Case report | 1/0 | 53 years | M | ASCT |
| **Balashov *et al.***<br>**(17)** | 2021 | Russia | Juvenile<br>myelomonocytic<br>leukemia | Case report | 1/0 | 9 months | F | HSCT |
| **Biernat *et al.***<br>**(18)** | 2021 | Poland | Haematological<br>malignancy (Acute<br>leukaemia/MDS,<br>CLL, Aggressive<br>Lymphoma, MM) | Retrospective<br>cohort study | 23/22 | NA | 62% M,<br>38% F |  |
| **Çınar *et al.***<br>**(19)** | 2020 | Turkey | MDS complicated<br>by recently<br>disseminated<br>tuberculosis with<br>associated kidney<br>disease | Case report | 1/0 | 55 years | M |  |
| **Dell'Isola *et al.* (20)** | 2021 | Italy | B-cell ALL | Case report | 1/0 | 6 years | F |  |
| **Ferrari *et al.* (21)** | 2021 | Italy | FL | Case report | 7/0 | 48 years | F |  |
|  |  |  | FL |  |  | 60 years | M |  |
|  |  |  | Indolent NHL |  |  | 60 years | M | SCT |
|  |  |  | Primary myelofibrosis |  |  | 43 years | M |  |
|  |  |  | DLBL |  |  | 70 years | M |  |
|  |  |  | ALL |  |  | 69 years | M | SCT |
|  |  |  | CLL |  |  | 60 years | M |  |
| **Hueso *et al.* (22)** | 2020 | France | B-cell lymphopenia | Observational multicentre study | 15/0 | 58 (35-77) | 5F, 12M |  |
| **Jeyaraman *et al.* (23)** | 2021 | India | Haematological malignancies | Retrospective observational multicentre study | 33/0 | 62 years (18–80) | 23M, 10F | 1 ASCT |
| **Karatas *et al.* (24)** | 2020 | Turkey | Mixed cellularity classical Hodgkin lymphoma and peripheral T-cell lymphoma | Case report | 1/0 | 61 years | M | ASCT |

CPT: convalescent plasma treated, CLL: chronic lymphocytic leukaemia, MML: myelomonocytic leukaemia, MM: multiple myeloma, HIV: human immunodeficiency virus, DLBL: diffused large B-cell lymphoma, NHL: non-Hodgkin's lymphoma, ALL: acute lymphocytic leukaemia, FL: follicular lymphoma. SCT: stem cell transplant, ASCT: autologous stem cell transplant, HSCT: haematopoietic stem cell transplant, MDS: Myelodysplastic syndrome, M: male, F: female.

### Data extraction and synthesis

The following clinical and laboratory variables were extracted: country, gender, age, type of haematological malignancy, cancer treatment, number of patients with CPT intervention, number of controls without CPT, other treatments for COVID-19. Clinical outcomes including, survival rate, adverse effect of CPT, the titter of antibody for donor and patient were recorded. Data analysis was qualitative, where the collected data were interpreted based on the presence or absence of the clinical outcomes. The findings were collected using Microsoft Excel sheets.

## Results

### Search findings

The outcome of the database search is described in a PRISMA flow chart (Figure 1). The search process yielded 2553 records, of which 578 articles were identified as duplicates and removed. Following title and abstract screening of the remaining 1975 articles, 1787 were excluded from the analysis due to one or more of the following reasons: 1) the article was published in a language other than English, 2) not original research article, 3) an animal-based study, 4) the full-text was not available, or 5) the CPT was not used as a COVID-19 treatment. From the 188 full texts screened, 171 studies were excluded because they were not conducted on patients with haematological malignancies. Seventeen studies met the inclusion criteria and were used to perform this systematic review. We identified 1103 patients, of which 258 patients had one or more haematological malignancy, diagnosed with COVID-19, and treated with CPT, whereas 845 patients were included in the control group and were provided with standard care in two of the identified studies (13, 18).

**Figure 1.**
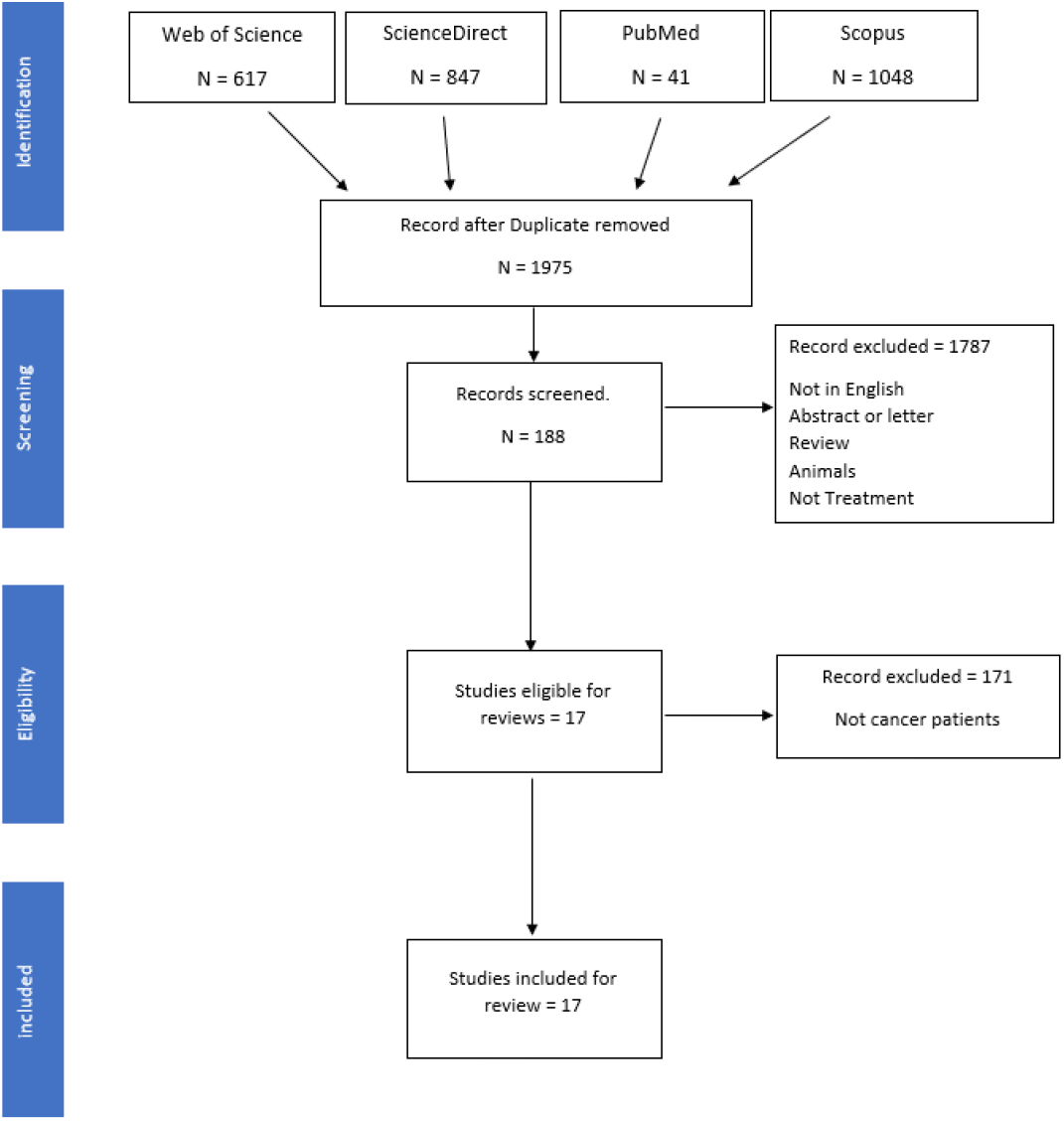
PRISMA Flow chart of study selection.

### Study characteristics and patients’ demographics

Among the 17 selected articles, 13 were case reports or case series, two were retrospective cohort studies, and two were observational multicentre studies (Table 1). The reported haematological malignancies were mainly follicular lymphoma (*n*=4), chronic lymphocytic leukaemia ((*n*=3), non-Hodgkin’s lymphoma (*n*=3), diffused large B-cell lymphoma (*n*=3), B-cell lymphoma (*n*=3), and unspecified haematological malignancies (*n*=4), (Figure 2). The range of patients age was from 9 months to 72 years, majority were older than 50 years while only three children were included (9 months, 4 and 5 years). A higher proportion of male patients were included in the studies compared to females (8:3). Ten patients had a history of receiving stem cell transplant, of which 7 were autologous transplants.

**Figure 2.**
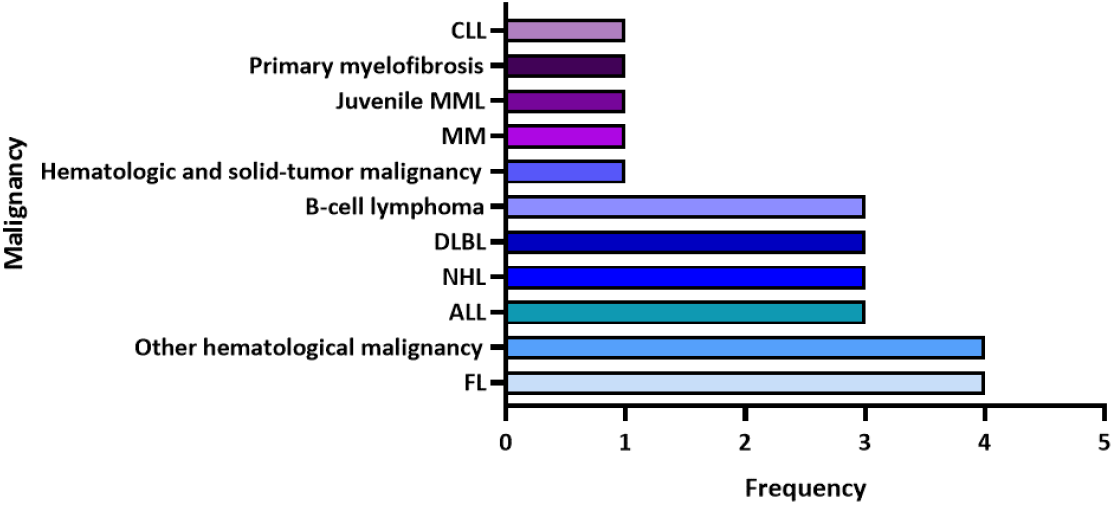
The frequency of haematological malignancies reported in the identified articles. CLL: chronic lymphocytic leukaemia, MML: myelomonocytic leukaemia, MM: multiple myeloma, HIV: human immunodeficiency virus, DLBL: diffused large B-cell lymphoma, NHL: non-Hodgkin’s lymphoma, ALL: acute lymphocytic leukaemia, FL: follicular lymphoma.

### Dose, time of administration and clinical outcomes of CPT

The dose of CP varied from 200 - 300 mL per transfusion. CPT was administered at variable time-points post hospital admission, where some patients received CPT as early as day 2-post diagnosis with COVID-19, while most of the remaining patients received CPT as a last treatment option after approximately one month of hospitalization (Table 3). In 14 studies, the total number of administered CPT doses ranged between 1-3 doses, however, two 53-year old patients diagnosed with lymphoma were administered 8 and 12 doses (2, 16) due to their poor response to the initial treatment.

**Table 3.** Summary of clinical outcomes from selected articles.

| Authors | Sample sizes<br>CPT/Non-CPT | Day of CPT<br>administration | No. of CP units<br>(dosage) | Outcome<br>endpoint | Outcome | Mortality | Adverse<br>events to<br>CPT | Drugs |
| --- | --- | --- | --- | --- | --- | --- | --- | --- |
| **Shankar et al. (6)** | 1/0 | Day 8 and 9 post illness onset | 2 (15 mL/kg) | 14 Days | Asymptomatic after 14 days | 0% | None | Oxygen therapy, Steroids (hydrocortisone and dexamethasone) |
| **Szwebel et al. (11)** | 1/0 | Day 65 and 66 post symptoms onset | 2 units daily | 70 Days | Asymptomatic after 70 days | 0% | None | Oxygen therapy, dexamethasone, oral prednisone, lopinavir/ritonavir, tocilizumab |
| **Wright et al. (12)** | 1/0 | NA | 1 (200 mL) | 1 month | Asymptomatic after 1 month and improvement of bilateral pulmonary infiltrates | 0% | None | Azithromycin and HCQ, oxygen therapy and supportive care |
| **Tremblay et al. (7)** | 24/0 | Median time: 3 days between doses | 2 (250 mL) |  | 13 patients were discharged home, 1 patient still hospitalized, and 10 patients died | 41.7% | 3 patients had FNHTR | Oxygen therapy and HCQ or azithromycin or remdesivir or tocilizumab or combination |
| **Thompson et al. (13)** | 143/823 | NA | - |  | Significantly improved 30-day mortality | 13.3% vs. 24.8% | None | Corticosteroids, remdesivir, tocilizumab, and HCQ |
| **Luetkens et al. (14)** | 1/0 | NA | 1 (200 mL) | 6 days | Asymptomatic at approximately 6 days from onset | 0% | None | Oxygen therapy |
| **Moore et al. (15)** | 1/0 | Day 88 post illness onset | 1 (200 mL) | 97 days | Asymptomatic at approximately 97 days from onset | 0% | None | Metoprolol for heart rate regulation and apixaban for anticoagulation |
| **Malsy et al. (16)** | 1/0 | Day 85 post illness onset | Two-course of 6 units (2 units/day administered every other day) | 140 days | Asymptomatic at approximately 140 days from onset | 0% | None | Remdesivir |
| **Rnjak et al. (2)** | 1/0 | Day 48, 49, 54, 55, 56, 57, 105 and 109 post illness onset | 8 (~200 mL) | 129 days | Afebrile with regression of pneumonia at 129 days from onset | 0% | None | Oxygen therapy, remdesivir and steroids |
| **Balashov et al. (17)** | 1/0 | Day 146 post HSCT | 3 (10 mL/kg) | 4 months | Complete viral clearance, full resolution of the lung lesions on CT | 0% | None | Tocilizumab and methylprednisolone |
| **Biernat et al. (18)** | 23/22 | Day 2-3 after diagnosis | 1-2 (200–250 mL) | Day 14 | Milder infection, less severe and faster resolution of symptoms, viral clearance | 13.0% vs. 41.0% | None | Oxygen therapy, mechanical ventilation, HCQ, Dexamethasone, Remdesivir, Tocilizumab, Lopinavir/Ritonavir |
| **Çınar et al. (19)** | 1/0 | Day 5 post symptoms onset | 2 (200 mL) | Day 7 | Improved dyspnea and fever resolution, viral clearance | 0% | None | Tocilizumab and favipiravir |
| **Dell'Isola et al. (20)** | 1/0 | Day 10 post admission | 3 (10 mL/kg) | Day 18 | Viral clearance | 0% | None | Remdesivir and prednisone |
| **Ferrari et al. (21)** | 7/0 | NA | 3 (210 mL) | Day 2 | COVID-19 symptoms resolved, viral clearance, radiological improvement | 0% | None | Corticosteroid, HCQ, low-molecular-weight heparin, and antibiotics |
|  |  | NA |  | Day 2 | COVID-19 symptoms resolved, viral clearance, radiological improvement |  |  | Corticosteroid, HCQ, low-molecular-weight heparin, and antibiotics |
|  |  | NA |  | Day 3 | COVID-19 symptoms resolved, radiological improvement |  |  | Corticosteroid, HCQ, low-molecular-weight heparin and antibiotics |
|  |  | NA |  | Day 7 | COVID-19 symptoms resolved, viral clearance |  |  | Corticosteroid, HCQ, low-molecular-weight heparin, and antibiotics |
|  |  | NA |  | N/A | COVID-19 symptoms resolved, viral clearance, radiological improvement |  |  | Corticosteroid, HCQ, low-molecular-weight heparin, and antibiotics |
|  |  | NA |  | Day 7 | COVID-19 symptoms resolved, viral clearance, radiological improvement |  |  | Corticosteroid, HCQ, low-molecular-weight heparin, and antibiotics |
|  |  | NA |  | Day 7 | COVID-19 symptoms resolved, viral clearance, radiological improvement |  |  | Corticosteroid, HCQ, low-molecular-weight heparin, and antibiotics |
| **Hueso et al. (22)** | 15/0 | Day 0 + 1 (2units each) | 4 (200-220 mL) | Day 2 | Fever resolved, and COVID-19 symptoms resolved after 2 weeks, decrease in RNAemia within 7-14 days | 5.9% | None | Remdesivir and tocilizumab |
| **Jeyaraman et al. (23)** | 33/0 | 4 days apart (range: 2–25 days) | 1-2 (200 mL) | Day 3 | Fever resolved | 45.5% | None | HCQ, remdesivir, favipiravir, other broad-spectrum antibiotics, steroids (methylprednisolone or dexamethasone), tocilizumab and oxygen support |
| **Karatas et al. (24)** | 1/0 | Day 40 post admission | 1 | Day 34 | Persistent SARS- CoV-2 viral shedding for 74 days | 0% | None | HCQ and azithromycin |

CPT: convalescent plasma treated, HCQ: Hydroxychloroquine.s

Different clinical outcome parameters were assessed in patients following CPT, but the most common reported outcomes were mortality, development of adverse events to CPT, clearance of SARS-CoV-2 infection, and cease of COVID-19 symptoms. Mortality was reported in 56 (21.7%) patients who received CPT and in 213 (25.2%) control patients who received the standard care. Adverse events to CPT were reported in only three patients who developed febrile non-haemolytic transfusion reaction. Other treatments that were administered to patients along with CPT including oxygen therapy, steroids, Azithromycin, Hydroxychloroquine, and Remdesivir.

The use of CPT is associated with improved overall survival (OR: 1.41, 95%CI: 0.99-1.99), viral clearance, (OR: 2.0, 95%CI: 1.04-2.08), detection of anti-SARS-CoV-2 antibodies in the patient’s plasma post CPT (OR: 6.33 95%CI: 1.7-17.3), hospital discharge (OR: 3.0 95%CI: 1.3-5.6), and recovery after 1 month of CPT (OR: 1.75 95%CI: 1.1-2.8). Further, the probability of developing adverse event in these patients due to CPT were significantly lower compared to the control group (OR: 0.24 95%CI: 0.14-0.40) (Figure 3).

**Figure 3.**
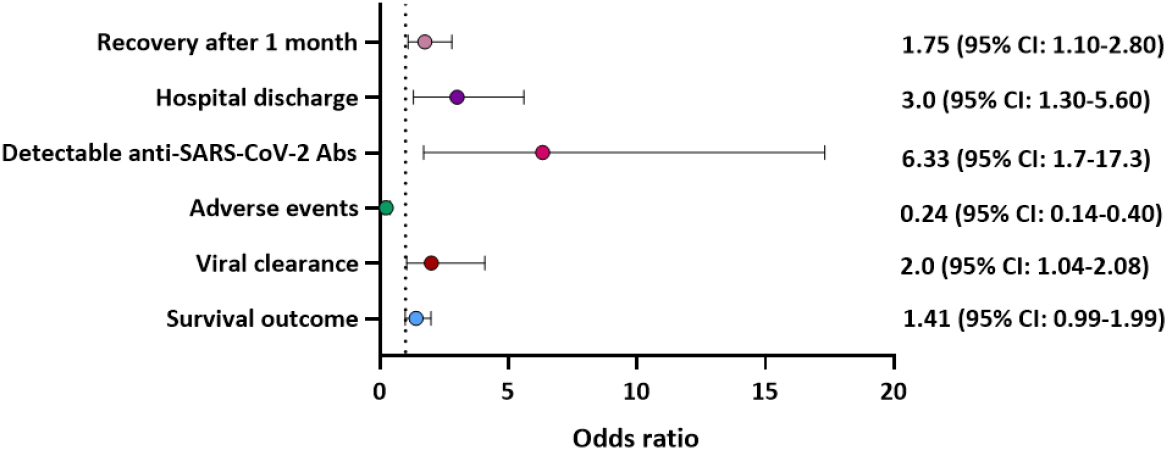
Odds ratio for the effectiveness of CPT on clinical outcome of patients with haematological malignancies and diagnosed with COVID-19.

## Discussion

Our analysis of the efficacy of using CPT from previously infected COVID-19 donors to treat COVID-19 infected patients with haematological malignancy suggests that CPT may reduce mortality rate and improve viral clearance.

A study by Borah *et al*. (25) reported a 60% mortality rate in haematological malignancies patients with severe COVID-19 infection. The efficacy of CPT to reduce the mortality rate was reported in four of the 17 studies, however, only two studies compared the percentage of mortality to the control group. Thompson *et al*. reported a mortality rate of 13.1% in the CPT recipient group and 24.8% in the non-recipient group, moreover, the mortality rate was significantly lower in intensive care unit patients on mechanical ventilation (26). Furthermore, another study demonstrated a mortality rate of 13% among the CPT recipient group compared to a mortality rate of 41% in the non-recipient group.

(27).

A recent meta-analysis reported that using CPT could significantly reduce the risk of mortality in covid-19 patients compare to those without CPT(28). Jeyaraman *et al*. (23)and Tremblay *et al*. (7) reported a mortality rate of 45.5% and 35.7%, respectively. However, the efficacy of CPT to reduce mortality in these two studies is difficult to ascertain due to the lack of a control group. Abeldaño Zuñiga *et al*. (29) concluded that using CPT for hospitalized COVID-19 patients could result in clinical improvement for patients, however, it does not significantly lower mortality rate in comparison to standard care and placebo.

Most of the included articles in our analysis reported significant improvement in patient status after CPT, patients become asymptomatic and were discharged. Four articles reported improved viral clearance after using CPT (17-20). Similarly, Sarkar *et al*. (30) concluded from 7 studies that CPT can enhance viral clearance and improve patient outcomes. on the other hand, a meta-analysis that included 5 RCTs, 6 cohort studies, and 16 case studies reported that using CPT in moderate and severe hospitalisedCOVID-19 patients does not affect the clinical outcome (31). However, in this meta-analysis the patients did not present with haematological malignanices and so they could have produced anti-SARS-CoV-2 antibodies, hence CPT intervention did not significantly improve their outcome.

In all reviewed articles, we found that CPT is safe with no adverse reactions reported in patients with haematological\ malignancies presenting with severe COVID-19 infection. However, one of the included studies reported febrile non-haemolytic transfusion reaction (FNHTR) in three patients post CPT (7). Supporting our findings, a recent mata-anlysis concluded that CPT is a safe approach for COVID-19 patient therapy where only 3.5% of recipients patients had developed an adverse effect (32). Nevertheless, a living systematic review stated that it is difficult to determine whether the adverse effect is due to the patient’s condition or CPT (33). Therefore, more studies are required to investigate the safety of CPT in COVID-19 infected cancer patients.

A minimum of one ABO compatible CP dose was given to patients, with most received 1 to 2 doses. However, one patient with diffuse large B-cell lymphoma (DLBCL) who had a prolonged active COVID-19 infection for 129 days, received 8 doses of CP until remission (34). There are no recommended or standardized doses of CP, however, most of the studies used one to two-units and the titer of the antibody might determine the optimal dose (35). In our study, only four papers measured the titer of the donor antibodies before transfusion (7, 17, 20, 24). One of the included retrospective multicentre observational studies found that there was no significant differences in moratlity in patients who received one versus two-dose of CP or in patients who received early versus late transfusion of CP (23).

In all 17 studies, standard care including supportive care and antiviral therapy with hydroxychloroquine, azithromycin, and/or ritonavir, and Tocilizumab (for very critical patients) were given to patients before initiating CPT. In addition, most of the patients required oxygen therapy. Duan et al. reported that CPT combined with supportive care and antiviral therapy can improve the clinical outcome of COVID-19 patients (36). Additionally, Agarwal *et al*. reported that using CPT alone might not be effective in reducing the severity, risk of mortality, and period of hospitalization of COVID-19 patients (37). Therefore, it is recommended to use CPT therapy with the combination of treatment to achieve the optimal result.

To the best of our knowledge, this is the first systematic review to discuss the efficacy and safety of CPT in patients with haematological malignancies presenting with severe COVID-19 complications. However, this review includes some limitations, including heterogeneity, lack of control group in most of the included studies, and randomized control trials. The heterogeneity of CPT dose, time of transfusion, reported outcome such as viral clearance, donor antibody titters, and lack of control groups to compare the result made it difficult to preform a meta-analysis.

## Conclusion

This review demonstrates that CPT is an effective and safe supportive therapy for patients with haematological malignancies and diagnosed with COVID-19 infection. The exact mechanism by which CPT may have mediated improved outcomes in the treated patients is likely multifactorial and could include reduction in viral load via enhanced clearance. Further studies and analysis are needed to fully understand the effectiveness of CPT in cancer patients with COVID-19.

## Data Availability

All data is available within the manuscript

